# Low-versus conventional-dose trimethoprim-sulfamethoxazole for non-HIV PCP

**DOI:** 10.1101/2023.04.14.23288508

**Authors:** Tatsuya Nagai, Hiroki Matsui, Haruka Fujioka, Yuya Homma, Ayumu Otsuki, Hiroyuki Ito, Shinichiro Ohmura, Toshiaki Miyamoto, Daisuke Shichi, Watari Tomohisa, Yoshihito Otsuka, Kei Nakashima

**Author notes:** Corresponding author: Kei Nakashima, Department of Pulmonology, Kameda Medical Center, 929 Higashi-cho, Kamogawa, Chiba 296-8602, Japan. **Take home message** Low-dose TMP-SMX treatment for non-HIV PCP showed no difference in mortality and fewer side effects compared with treatment with conventional doses. Therefore, it may be an effective alternative, but further research is needed to confirm our results.

## Abstract

Trimethoprim-sulfamethoxazole (TMP-SMX) is an effective treatment for *Pneumocystis jirovecii* pneumonia (PCP); however, a high incidence of adverse events has been observed. Low-dose TMP-SMX is a potentially effective treatment with fewer adverse events; however, evidence is limited. We aimed to evaluate the efficacy and safety of low-dose TMP-SMX after adjusting for patient background characteristics.

In this multicentre retrospective cohort study, we included patients diagnosed with non-human immunodeficiency virus (HIV) PCP and treated with TMP-SMX between June 2006 and March 2021 at three institutions. The patients were classified into low-(TMP equivalent <12.5 mg/kg) and conventional-dose groups (TMP equivalent 12.5–20 mg/kg/day). The primary endpoint was 30-day mortality, and the secondary endpoints were 180-day mortality, adverse events of grade 3 or greater per the Common Terminology Criteria for Adverse Events Version 5.0, and initial treatment completion rates. The background characteristics were adjusted using the overlap weighting method with propensity scores.

Fifty-five patients in the low-dose and 81 in the conventional-dose groups were evaluated. There was no significant difference in 30-day mortality (7.6% vs. 14.9%, *P* = 0.215) or 180-day mortality (18.1% vs. 24.0%, *P* = 0.416) after adjusting for patient background characteristics. The incidence of adverse events, especially nausea and hyponatremia, was significantly lower in the low-dose group (27.3% vs. 58.6%, *P* = 0.001). The initial treatment completion rates were 43.8% and 27.7% in the low-dose and conventional-dose groups, respectively.

Low-dose TMP-SMX did not alter survival but reduced the incidence of adverse events in patients with non-HIV PCP, compared with conventional-dose TMP-SMX.

## Introduction

*Pneumocystis jirovecii* pneumonia (PCP) is a widely known opportunistic infection in immunocompromised patients [1, 2]. The occurrence of PCP in patients with acquired immune deficiency syndrome has been problematic worldwide since the 1980s; however, the frequency of PCP in this population has decreased significantly owing to early treatment for human immunodeficiency virus (HIV) in recent years [2, 3]. Nevertheless, the occurrence of PCP in non-HIV patients has increased owing to the use of immunosuppressive drugs, anticancer drugs, and the increasing number of patients undergoing organ transplantations [2, 4]. Furthermore, the mortality rate (30–60%) of non-HIV PCP is higher than that (10–20%) of HIV PCP owing to a strong inflammatory response to a smaller number of organisms, unlike milder immune reactions in HIV PCP [1, 2]. Therefore, it is crucial to improve the therapeutic management of PCP in non-HIV immunocompromised patients to improve their prognoses.

Prior reviews and guidelines indicate that the first-line therapy for patients with non-HIV PCP and HIV PCP is trimethoprim-sulfamethoxazole (TMP/SMX), and doses of 15–20 mg/kg/day TMP and 75–100 mg/kg/day SMX are recommended [1, 2, 5-8]. The efficacy of TMP/SMX as a standard therapy is clear; however, it is associated with a high incidence of adverse events, including drug rashes and gastrointestinal, liver, dose-dependent renal, and blood disorders, that make it difficult to continue treatment [9-11]. Therefore, studies on low-dose therapy (TMP 10–15 mg/kg/day or less) and step-down therapy with TMP/SMX have been conducted; these suggested comparable therapeutic effects and the reduction of the incidence of adverse effects compared with that in standard therapy [11-16]. However, the evidence is insufficient because there are only a small number of retrospective studies [11-15], and the patient background between the low-dose and conventional-dose groups was not adjusted to compare mortality rates and safety [16]. In a prior study, Hammarstrom et al. found no differences in respiratory function changes between the baseline and day 8 in low-dose and conventional-dose TMP-SMX groups by employing an adjusted regression model for treating PCP patients with haematological malignancies [16]. However, this study did not evaluate 30-day mortality or adverse reactions after adjusting for potential confounders, and it was limited to patients with haematological malignancies as their underlying diseases.

Therefore, we aimed to evaluate the efficacy and safety of low-dose TMP/SMX compared with those of conventional-dose TMP/SMX in non-HIV PCP patients with various underlying diseases. We set 30-day mortality as the primary endpoint after adjusting for patient background in a multicentre cohort study.

## Methods

### Study Design and Study Population

This was a multicentre retrospective observational cohort study involving clinically diagnosed non-HIV PCP patients treated with TMP-SMX. We retrospectively enrolled non-HIV patients diagnosed with pneumocystis pneumonia between January 2006 and March 2021 at Kameda General Hospital, Seirei Hamamatsu Hospital, and Seirei Mikatahara Hospital (registry named RE-VISION-PCP: Registry to Provide New Evidence and Insights for the Management of Pneumocystis Pneumonia in Non-HIV-infected Patients). The diagnostic criteria for PCP were defined as follows [11, 17]: 1) clinical symptoms consistent with those of PCP (dyspnoea, cough, fever, hypoxemia), 2) chest X-ray or chest CT scans showing abnormal shadows (such as bilateral ground-glass opacities) consistent with PCP findings, 3) *Pneumocystis* detected in respiratory specimens such as sputum or bronchoalveolar lavage fluid through conventional staining (Grocott methenamine silver or Diff-Quick staining) or deoxyribonucleic acid testing (loop-mediated isothermal amplification or polymerase chain reaction), and 4) elevated β-D-glucan concentrations with an appropriate response to standard therapy for PCP. Serum β-D-glucan concentrations were measured using either the β-D-glucan test kit (Wako Pure Chemical Industries, Osaka, Japan) or the FUNGITEC G test MKII (Nissui Pharmaceutical, Tokyo, Japan). Elevated β-D-glucan levels were defined as >5 pg/mL (β-D-glucan test; Wako assay) or >20 pg/mL (FUNGITEC G-test KM assay) [18, 19]. 1) Patients who did not receive any treatment, 2) Patients who were initially treated with regimens other than TMP/SMX, and 3) patients receiving >20 mg/kg of TMP were excluded. The study protocol was reviewed and approved by the research ethics committees of Kameda General Hospital (#21-069), Seirei Hamamatsu Hospital (#3584), and Seirei Mikatahara Hospital (#21-58). This was a retrospective study, and the patient information was anonymized; therefore, written informed consent was not obtained.

### Definitions of the Low-Dose and Conventional-Dose TMP-SMX Treatment Groups

Patients were divided into low- and conventional-dose TMP-SMX treatment groups according to the amount of TMP-SMX. The dosage of TMP-SMX was calculated by multiplying a correction factor according to creatinine clearance because dose adjustment and reduction according to the degree of renal dysfunction are necessary [13]. Creatinine clearance was calculated using the Cockcroft–Gault formula [20]. The specific correction coefficients were classified as 1 for CrCl > 50 ml/min; 1.5 for CrCl 30−50 mL/min; 2.0 for CrCl 15−30 mL/min; and 3.0 for CrCl < 15 mL/min or haemodialysis use [13]. The low-dose group was defined as patients treated with an initial dose of TMP <12.5 mg/kg/day, while the conventional-dose group was defined as patients treated with an initial dose of TMP of 12.5−20 mg/kg/day. Previous studies have defined low-dose TMP-SMX as TMP at 10–15 mg/kg/day [11-16]. Additionally, a threshold of 13.85 mg/kg/day has been reported as the point of severe adverse reactions in the corresponding area under the receiver operating characteristic curves used to distinguish patients with severe adverse reactions [14]. Thus, we established a TMP dose of 12.5 mg/kg/day, which represents the median dose of 10–15 mg/kg/day, as the cut-off point for low-dose treatment.

### Outcomes

The primary endpoint was 30-day mortality from the start of treatment. The secondary endpoints were mortality at 180 days from the beginning of treatment, the incidence of each adverse event, and the completion rate of initial treatment.

### Data Collection

Data on patient characteristics including age, sex, weight, underlying diseases, laboratory results (platelet count; haemoglobin, serum albumin, lactate dehydrogenase, serum sodium, serum potassium, and creatinine levels; creatinine clearance), state of consciousness, presence of hypotension, respiratory status (including no oxygen support, oxygen support, and mechanical ventilation with options like high-flow nasal cannula, non-invasive positive-pressure ventilation, and intubation), dose of TMP-SMX, use of adjunctive glucocorticoid therapy (steroid pulse therapy is defined as the initial administration of methylprednisolone at 500mg-1000mg/day, while a mild to moderate doses is defined as a steroid dose less than that of steroid pulse therapy), 30- and 180-day mortality rates, the occurrence of grade 3 or higher adverse events, and the discontinuation of the initial regimen were collected retrospectively from medical records. Adverse drug reactions were evaluated according to the Common Terminology Criteria for Adverse Events Version 5.0 [21]; this included grade 3 or higher skin rashes, nausea, decreases in white blood cell counts, anaemia, thrombocytopenia, increases in plasma aspartate aminotransferase (AST) levels, increases in plasma alanine aminotransferase (ALT) levels, hyponatremia, and hyperkalaemia.

### Statistical Analyses

Because of the retrospective and observational nature of this study, we conducted it using the available number of cases and did not perform any sample size calculations. We obtained descriptive statistics of the baseline characteristics and outcomes between the low- and conventional-dose groups. No missing values were observed in the variables included in the analysis. Continuous variables are expressed as mean ± standard deviation, and categorical variables are expressed as numbers (%). The balance between the groups for each baseline characteristic was assessed using the standardized mean difference (SMD). Generally, groups are considered balanced when the SMD is less than 0.1 [22]. Outcomes between the groups were tested using *t*-tests for continuous and the χ-square test for categorical variables. The significance level was set to 0.05. We used overlap weighting with a propensity score to obtain risk-adjusted cohorts [23]. Propensity scores were calculated using logistic regression with the exposure as the dependent variable and age, sex, history of malignancies, serum albumin levels, lactate dehydrogenase levels and respiratory status as the explanatory variables. These explanatory variables are prognostic factors for pneumocystis pneumonia [5, 24]. We described the patient demographics of the risk-adjusted cohorts similarly to those of the unadjusted cohort. We evaluated the balance between the groups using the SMD. Outcomes were also compared between the groups similarly to those of the unadjusted cohort.

## Results

A flowchart of the patient selection process is shown in Figure 1. In total, 164 non-HIV immunocompromised patients diagnosed with PCP were included. Among them, five patients who did not receive any treatment, 16 who received initial treatment other than TMP-SMX, and seven who received more than 20 mg/kg of TMP were excluded. Finally, 81 patients were included in the conventional-dose group and 55 in the low-dose group.

**Figure 1.**
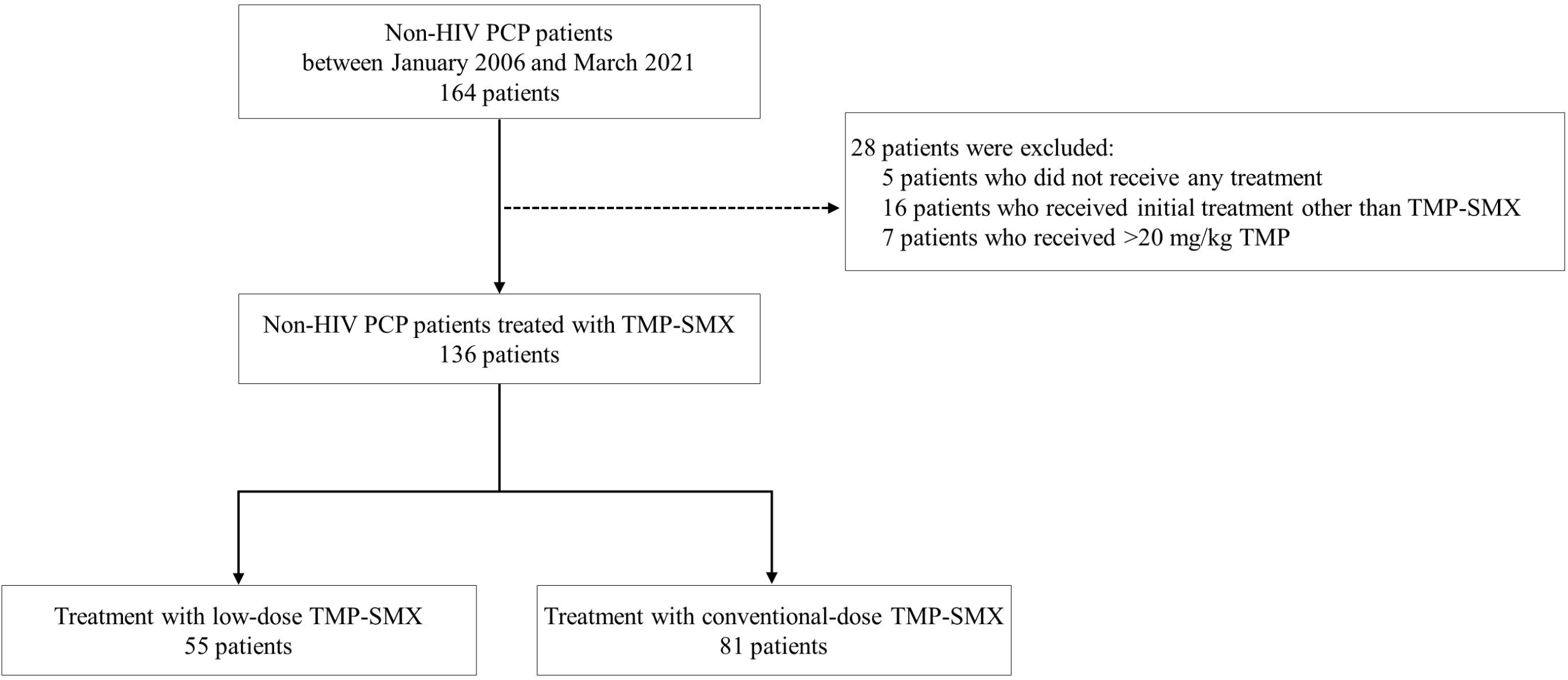
Patient selection flowchart HIV, human immunodeficiency virus; TMP-SMX, trimethoprim-sulfamethoxazole; PCP, *Pneumocystis jirovecii* pneumonia

The demographic and clinical unadjusted and adjusted characteristics of eligible patients stratified by the TMP-SMX dose are summarized in Table 1. Patient characteristics with SMDs above 10% were unbalanced, while those with SMDs below 10% were balanced. Sex, median weight, and the presence of malignancies or connective tissue disease were unbalanced. Among the haematology findings, the platelet count was balanced, while the other parameters were unbalanced. Hypotension (systolic blood pressure <90 mmHg) and respiratory status were unbalanced, whereas glucocorticoid non-use was balanced. Adjusted patient characteristics included age, sex, underlying diseases, serum albumin levels, serum lactate dehydrogenase levels, and respiratory status, with SMDs less than 10%.

**Table 1.**
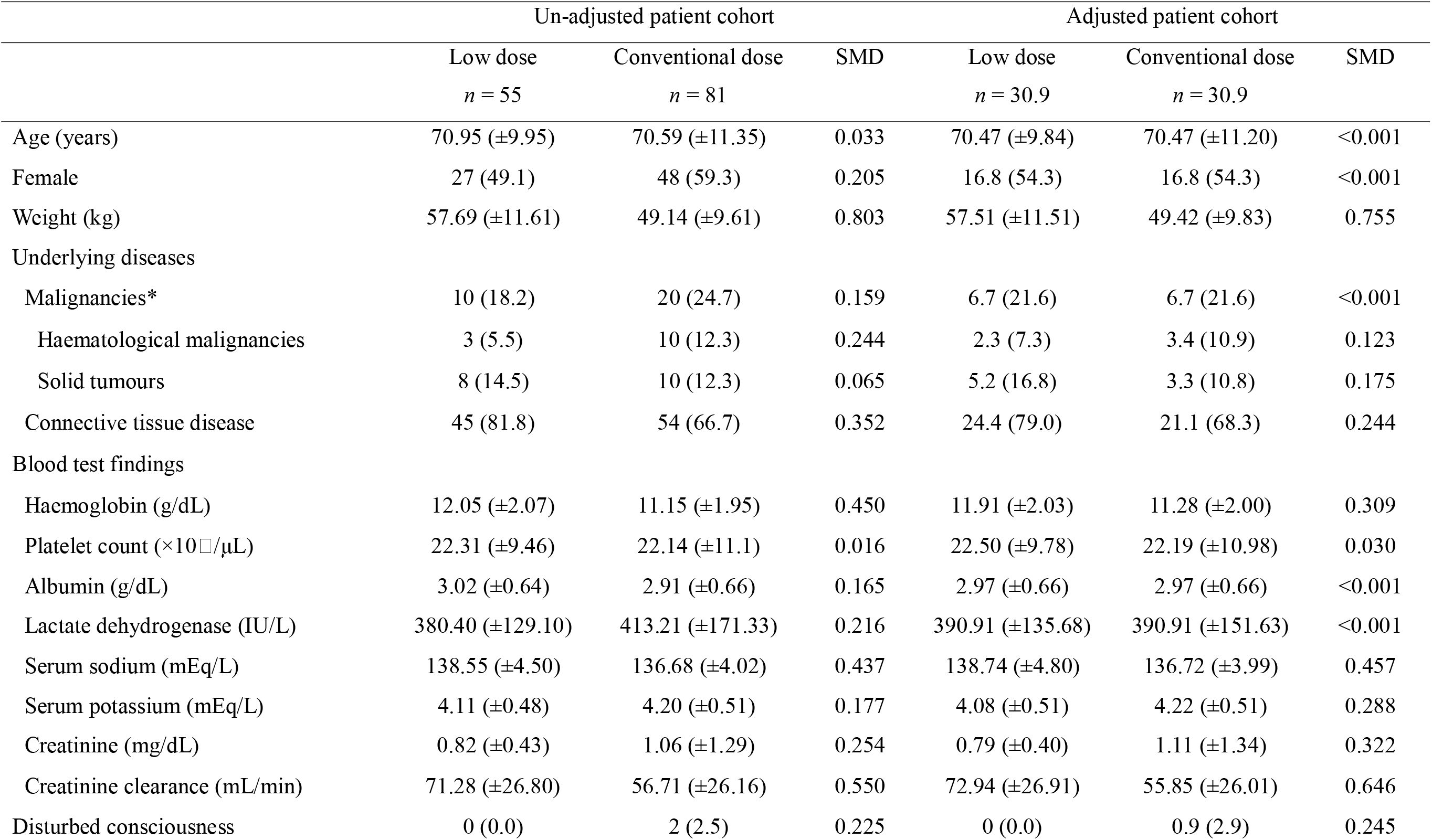
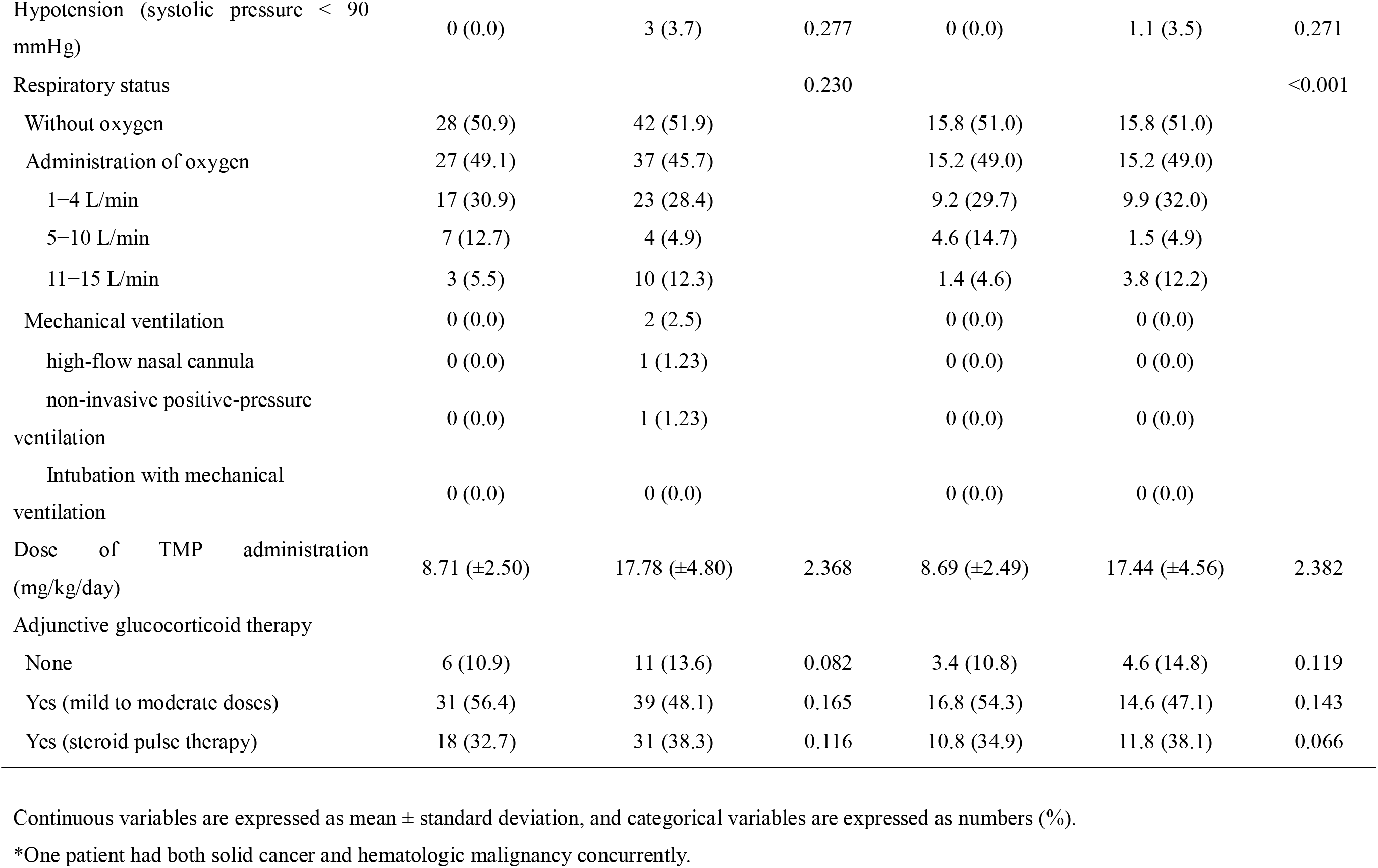
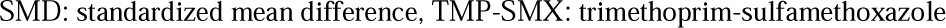
Baseline clinical characteristics of the study population (when administered with TMP/SMX)

Data on the primary and secondary endpoints for eligible patients stratified by the TMP-SMX dose are presented in Table 2. The unadjusted 30-day mortality rate was 7.3% in the low-dose group and 17.3% in the conventional-dose group. The 180-day survival rate was 18.2% in the low-dose group and 28.4% in the conventional-dose group; however, the difference was not significant. Similarly, the adjusted mortality rates were not significantly different (7.6% in the low-dose group and 14.9% in the conventional-dose group for 30-day mortality, and 18.1% in the low-dose group and 24.0% in the conventional-dose group for 180-day mortality). The unadjusted total adverse reaction rate was significantly lower in the low-dose group (29.1%) than in the conventional-dose group (56.8%). The adjusted total adverse reaction rate was also significantly lower in the low-dose group (27.3%) than in the conventional-dose group (58.6%). In the unadjusted population, the incidence of nausea grade 3 or higher was 0% in the low-dose group and 9.9% in the conventional-dose group and that of hyponatremia was 7.3% in the low-dose group and 28.4% in the conventional-dose group; the incidence of these adverse reactions was significantly lower in the low-dose group than in the conventional-dose group. The incidence of nausea and hyponatremia was lower in the low-dose group than in the conventional-dose group in the adjusted population. In contrast, there were no significant differences in the incidence of skin rashes, leukopenia, anaemia, thrombocytopenia, ALT elevation, or hyperkalaemia. The unadjusted initial treatment completion rate was 43.6% in the low-dose group and 28.4% in the conventional-dose group, while the adjusted initial treatment completion rate was 43.8% in the low-dose group and 27.7% in the conventional-dose group.

**Table 2.**
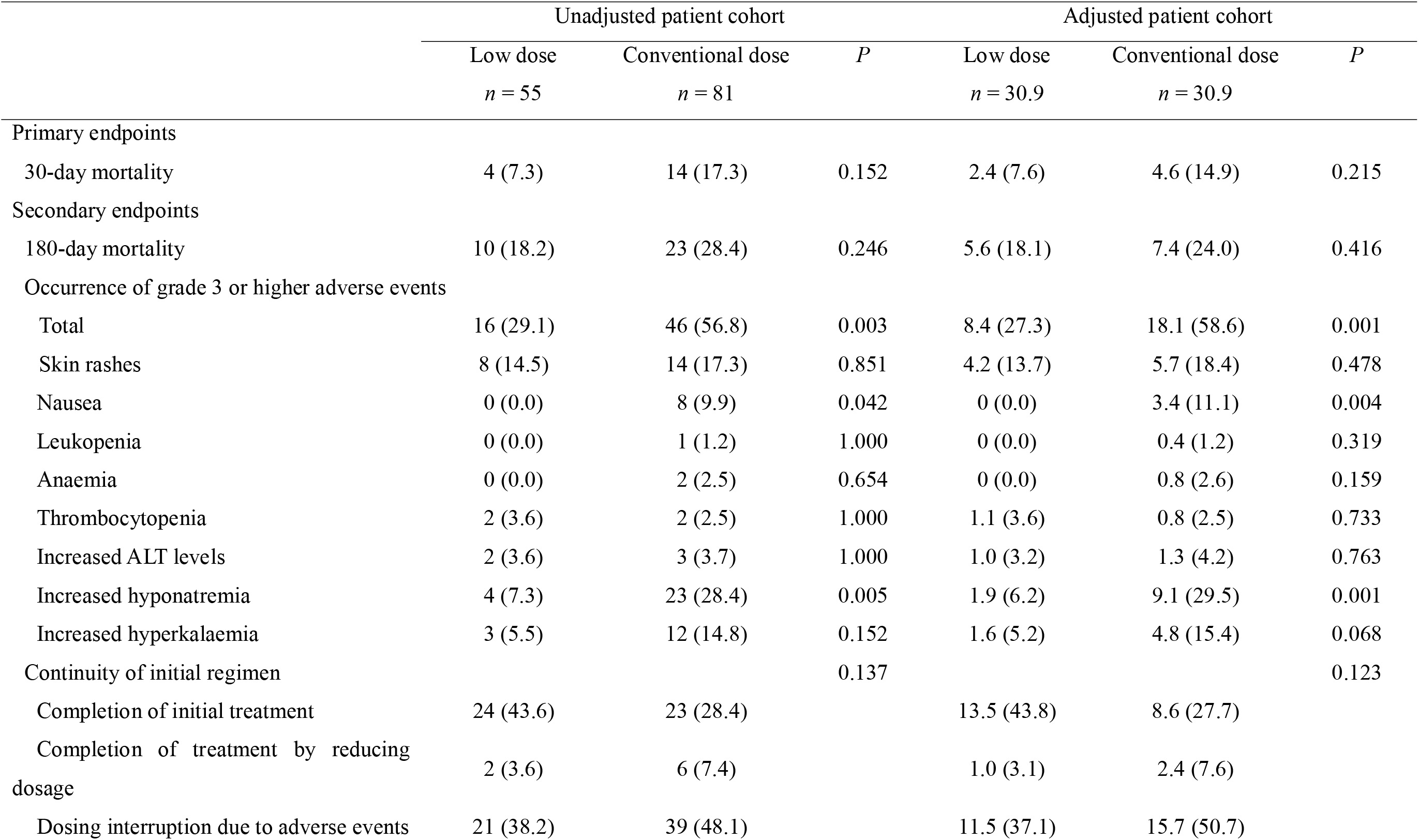
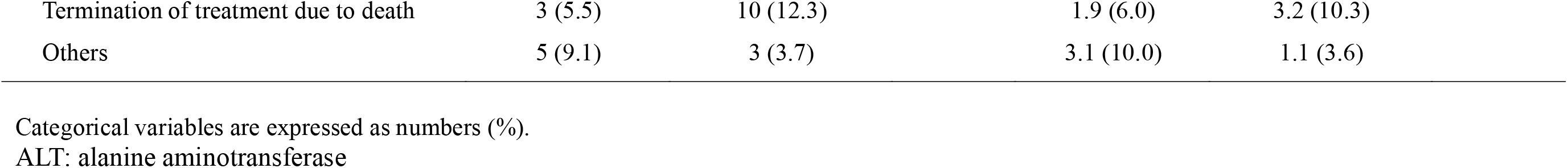
Outcome data

## Discussion

This multicentre retrospective observational study evaluated the treatment outcomes of low-dose versus conventional-dose TMP-SMX in patients with non-HIV PCP after adjusting for patient demographics and clinical characteristics. There were no significant differences in the 30- and 180-day survival rates between the low- and conventional-dose groups. However, the rate of total adverse events of grade ≥ 3 was significantly lower in the low-dose group than in the conventional-dose group. To the best of our knowledge, this is the first and largest multicentre study to evaluate the mortality rate and safety of low-dose TMP-SMX for non-HIV PCP after adjusting for patient characteristics. We believe that our findings offer additional evidence supporting the effectiveness of low-dose TMP-SMX treatment for the management of non-HIV PCP.

Recent guidelines set the conventional dose of TMP and SMX at 15–20 mg/kg and 75–100 mg/kg, respectively [5-7]. However, because of concerns about adverse events and tolerability of the conventional dose and because the current recommended dose for non-HIV PCP is based on small observational studies conducted in the 1970s and 1980s [25-27], several studies have recently examined the efficacy of low-dose or step-down treatment with TMP-SMX. In 2020, Butler-Laporte et al. published a systematic review and meta-analysis of low-dose TMP-SMX for PCP treatment; the treatment of PCP with trimethoprim at doses of 15 mg/kg/day or less had similar mortality rates and significantly fewer severe adverse events attributable to the treatment than did treatment with conventional doses [15]. Among the included studies, three [11, 13, 14] were retrospective studies of conventional and low doses, with mortality and adverse effects as outcomes. In one study (where a low dose was defined as TMP < 10 mg/kg/day), no comparisons were made, and it was purely descriptive [11]. Two of the studies comparing mortality and adverse events between low- and conventional-dose TMP-SMX for PCP showed no statistically significant difference in mortality [13, 14], although patient background was not adjusted for in either study. Recently, Hammarstrom et al. compared low-dose (TMP equivalent 7.5−15 mg/kg/day) and conventional-dose TMP-SMX (TMP equivalent 15−20 mg/kg/day) in patients with haematological malignancies [16]. They evaluated the change in respiratory function measured as the difference in PaO_2_/FiO_2_ ratio between the baseline and day 8 as a primary endpoint. There were no significant differences in the primary endpoint between the two groups, either before or after controlling for potential confounders in an adjusted regression model [16]. The target population for this study was restricted to patients with haematological malignancies, and only three covariates (age, Eastern Cooperative Oncology Group performance status, and P/F ratio at baseline) were utilized for adjustment. In contrast, our study evaluated the mortality and safety of low-dose TMP-SMX (TMP < 12.5 mg/kg/day) in non-HIV PCP patients with a wide range of underlying diseases, including collagen disease, solid tumours, and haematological malignancies. We adjusted for patient background by considering six prognostic factors that influence mortality in non-HIV PCP patients: age, sex, history of malignancies, serum albumin levels, serum lactate dehydrogenase levels and respiratory status [5, 24].

In the present study, the 30- and 180-day mortality rates of the low-dose group were 7.6 and 18.1%, respectively, after adjusting for patient background; they were not significantly different from those of the conventional-dose group. There are several possible explanations for this result. First, it is possible to obtain sufficient minimum inhibitory concentration (MIC) for *Pneumocystis jirovecii* in blood and tissues, even at low doses. To ensure adequate therapeutic efficacy of TMP-SMX, a dose above the MIC must be ensured [28]; however, continuous *in vitro* cultures of *Pneumocystis* for testing its susceptibility to antimicrobial agents have not been performed [29]. Therefore, the required dosage for PCP treatment was determined based on the results of clinical studies [11]. One tablet of TMP-SMX (80 mg of TMP and 400 mg of SMX; equivalent to 1.3 mg/kg/day TMP and 6.7 mg/kg/day SMX for a 60 kg body) once daily is recommended and is sufficient for preventing the onset of PCP [30]. Generally, the dose of antimicrobial agents required to prevent disease onset has a concentration above the MIC in many infections [31]. Therefore, lower doses of TMP-SMX than conventional-dose (TMP equivalent 15-20 mg/kg/day) could have a sufficient therapeutic effect [11]. Second, there were fewer adverse events in the low-dose group than in the conventional-dose group in the present study. TMP-SMX is associated with a dose-dependent incidence of adverse events, such as skin rashes, gastrointestinal disturbances, myelosuppression, renal disturbances, hepatic disturbances, and electrolyte disturbances [32]; 57% of patients with HIV PCP change from this treatment owing to serious adverse events [33]. In the current study, after adjusting for patient background, there were significantly fewer adverse events in the low-dose (27.3%) than those in the conventional-dose groups (58.6%), with nausea and hyponatremia being significantly less common. This reduction in the incidence of adverse events could have facilitated the management of PCP. Therefore, low-dose TMP-SMX may be an effective treatment for non-HIV PCP and should be considered a treatment option.

The definition of low-dose TMP-SMX treatment remains unclear, with some studies defining it as TMP < 10 mg/kg/day [11, 14] and others as TMP < 15 mg/kg/day [13, 16]. In a study by Ohmura et al., the optimal dosage of TMP/SMX treatment to minimize severe adverse events was investigated using receiver operating characteristic curves, reporting a threshold TMP dose of 13.8 mg/kg/day [14]. Consequently, in our study, we referred to this value and defined the low dose as TMP < 12.5 mg/kg/day. Further investigation is warranted to determine the optimal threshold for low-dose TMP-SMX combination therapy.

Our study findings suggest that low-dose TMP-SMX treatment may be a viable option for clinicians. However, future research, including large-scale observational studies and randomized controlled trials, is needed to confirm the efficacy of low-dose TMP-SMX.

This study has some limitations. First, this was a retrospective observational study. Therefore, there could be a bias in patient selection and treatment choice. We adjusted as much as possible for the mortality risk factors that could affect patient outcomes; however, there could have been unrecognized confounding factors. Second, the blood levels of TMP-SMX were not measured; therefore, it is unclear whether there is a true correlation between actual blood levels and treatment response or adverse events. Maintaining the blood levels of TMP-SMX in the appropriate range is important to ensure therapeutic efficacy and reduce the incidence of adverse events; however, actual blood levels are dependent on renal function, liver function, and body size [34]. Third, only a few patients with severe respiratory failure, such as those on ventilators, were included, and the efficacy of TMP-SMX in such patients with severe respiratory failure is unknown.

## Conclusions

Low-dose TMP-SMX treatment for non-HIV PCP was associated with no difference in mortality and few side effects compared with those of conventional-dose treatment. Low-dose TMP-SMX may be an effective treatment option for non-HIV-associated PCP. The retrospective observational nature of the study and the small number of seriously ill patients warrant further investigations in larger prospective randomized controlled trials or cohort studies.

## Data Availability

All data produced in the present study are available upon reasonable request to the authors

## Conflicts of interest

None.

### Acknowledgements

We thank the Department of Medical Oncology, Haematology/Oncology, and Rheumatology doctors at Kameda Medical Center for providing the case data.

## CRediT statement

Conceptualization: T.N., H.M., S.O., K.N. Data curation: T.N., H.M., S.O., K.N. Formal analysis: H.M. Investigation: T.N., H.F., Y.H., S.O., D.S. Methodology: T.N., H.M., S.O., K.N. Project administration: T.N., S.O., K.N. Resources: T.N., H.F., Y.H., A.O., H.I., S.O., T.M., D.S., W.T., Y.O., K.N. Software: H.M. Supervision: K.N. Validation: H.M., K.N. Visualization: T.N., H.M., K.N. Writing - original draft: T.N. Writing - review & editing: T.N., H.M., H.F., Y.H., A.O., H.I., S.O., T.M., D.S., W.T., Y.O., K.N.

